# LACK OF ASSOCIATION BETWEEN CONVALESCENT PLASMA ADMINISTRATION AND LENGTH OF HOSPITAL STAY: A HOSPITAL-DAY STRATIFIED MULTI-CENTER RETROSPECTIVE COHORT STUDY

**DOI:** 10.1101/2021.05.04.21256627

**Authors:** Joy Alamgir, M. Ruhul Abid, Brian Garibaldi, Naved Munir, Soko Setoguchi, Stephanie S. Hong, Xinci Chen, Paul T. Kocis, Masanao Yajima, G. Caleb Alexander, Hemalkumar B. Mehta, Vithal Madhira, Rosa Ergas, Thomas R. O’Brien, Sam Bozzette

**Author notes:** Correspondence Joy Alamgir, Principal Investigator, ARIScience, 148 Commonwealth Rd STE 5138, Wayland MA 01778, Ruhul Abid MD PhD, Associate Professor, Brown University Warren Alpert Medical School, Cardiothoracic Surgery, Rhode Island Hospital, 1 Hoppin St, Providence RI 02903.

## Abstract

**Background:** Effects of timing of Convalescent plasma (CP) administration on hospitalized COVID-19 patients are not established.

**Methods:** We used the National COVID Cohort Collaborative data to perform a retrospective cohort study of hospitalized COVID-19 patients in the United States between 07-01-2020 and 12-19-2020. We stratified patients based on day of CP administration (Day 0, 1, 2, 3 and 4) from COVID-19 diagnosis. We used 35 predictors to frame matched cohorts accounting for clinical and sociodemographic characteristics. We used competing risk survival models to examine the association between CP administration and length of hospital stay with in-hospital death as a competing risk performing Gray’s test on the cumulative incidence function and Cox’s regression on cause specific hazard ratios.

**Results:** In a cohort of 4,003 hospitalized COVID-19 patients, 197 (4.9%) received CP within the first 5 days following COVID-19 diagnosis. After adjusting for potential confounding variables, there were no statistically significant associations between day of CP administration and length of hospital stay. Day 0 CP administration signallled lower mortality but was not statistically significant (HR 0.45 [0.19-1.03]).

**Conclusions:** We found no association between the timing of CP administration and length of stay among hospitalized COVID-19 patients.

## INTRODUCTION

As of April 5, 2021, 554,064 individuals have died and more than 30.5M have been infected with SARS-CoV-2, the virus that causes COVID-19, in the United States.^1^ Given the morbidity and mortality that the pandemic has posed, an extraordinary effort has been undertaken to identify effective treatments for COVID-19 in addition to vaccination efforts^2^. Among potential candidates under active investigation, there has been continued interest in the potential value of convalescent plasma (CP) given its availability from those who have recovered from COVID-19 infection. Federal Drug Administration (FDA) issued an emergency use authorization (EUA) for CP as a treatment for hospitalized COVID-19 patients on August 23, 2020.^3^

To better understand the potential effects of CP as a treatment for COVID-19, there have been studies in multiple countries (e.g., US, Argentina, Bahrain, China, India, Spain, The Netherlands) involving different patient populations.^4-16^ These studies, ranging from RCTs to observational studies, have evaluated the reduction in mortality and length of hospitalization, as well as other secondary endpoints. However the effectiveness of CP administration has been mixed in these studies. For example Gharbharan et al in their RCT reported no improvement in mortality and that by the time COVID-19 patients are hospitalized there are already neutralizing antibodies in the patients at a level comparable to donor CP. Simonovich et al and Agarwal et al in their respective RCTs also reported no difference in outcome with CP administration. In contrast Libster et al in their RCT reported improvement in disease progression. Observational studies, with or without matching, also reported conflicting results. For example Bodhiraj et al, and Liu et al, and Joyner et al observed improved outcomes from CP administration while Rogers et al and Sostin et al did not. There are several plausible reasons for differences in the conclusions drawn by these varied studies, including differences in study populations, timing of CP exposure, outcome ascertainment and statistical methods used. It is noteworthy that while some studies looked at early and late CP administration effects (Salazar et al) we were unable to find any studies that did a per-day stratification of the effect of CP administration to identify the specific effects of the timing of CP administration. This research aims to fill the knowledge gap related to timing of CP administration and COVID-19 outcomes.

In this analysis, we tested the hypothesis that CP administration reduces the duration of hospitalization in COVID-19 patients and that this reduction is dependent on the day of administration (Day 0, 1, 2, 3 and 4) of COVID-19 diagnosis. We used the National COVID Cohort Collaborative (N3C)^17^ data to test the hypothesis.

## METHODS

### N3C Data

The N3C harmonizes Electronic Health Record data for COVID-19 tested persons dating from 01/01/2018. Data from 36 medical centers were securely transferred to the N3C data enclave and harmonized into a single common model.^18-19^ We included patients who were diagnosed and hospitalized with COVID-19 from July 1, 2020 to December 19, 2020. As of 12/19/2020, N3C contained 2.5M total patients including 400K COVID-19 patients, 1.13B lab results, 472M drug exposures, and 185M procedures. We froze our analysis on that release to minimize confounding from COVID-19 vaccination, which began in the United States in mid-December 2020.^20^

### Study Institutions and Patients

Figure 1 shows the population selection for this analysis and the distribution of day of CP administration as compared to COVID-19 diagnosis. Data-providers were excluded from the analysis if their data did not include specific codes or descriptions for CP data that allowed us to differentiate CP from other plasma products.^21^ Patients not diagnosed with COVID-19 were excluded. Patients for whom data on age or gender were missing were excluded. To focus on patients at higher risk of severe disease, we also excluded patients less than 30 years of age. Patients more than 85 years of age were excluded given the likelihood of other severe comorbidities that might have independently influenced their hospital stay and the possibility of death confounded by do-not-resuscitate, do-not intubate orders. We further restricted the analysis to patients who were hospitalized. To reduce confounding introduced by decreased COVID-19 mortality among hospitalized from March 2020 to July 2020, patients diagnosed with COVID-19 before July 1, 2020 were excluded.^22^ To eliminate the effect of COVID-19 vaccination in our analysis, we excluded patients diagnosed after December 19, 2020. We worked with N3C operational staff to identify a blacklist of patients from early in the COVID-19 pandemic. This blacklist methodology was required as our data access was de-identified with dates shifted +/-180 days. Thus effectively the time period of the cohort ranged from July 1, 2020 to December 19, 2020. Missing predictor data were handled using a constant value and separately a flag indicating that data were missing and as such were part of the propensity score calculation described below.^23^

**Figure 1:**
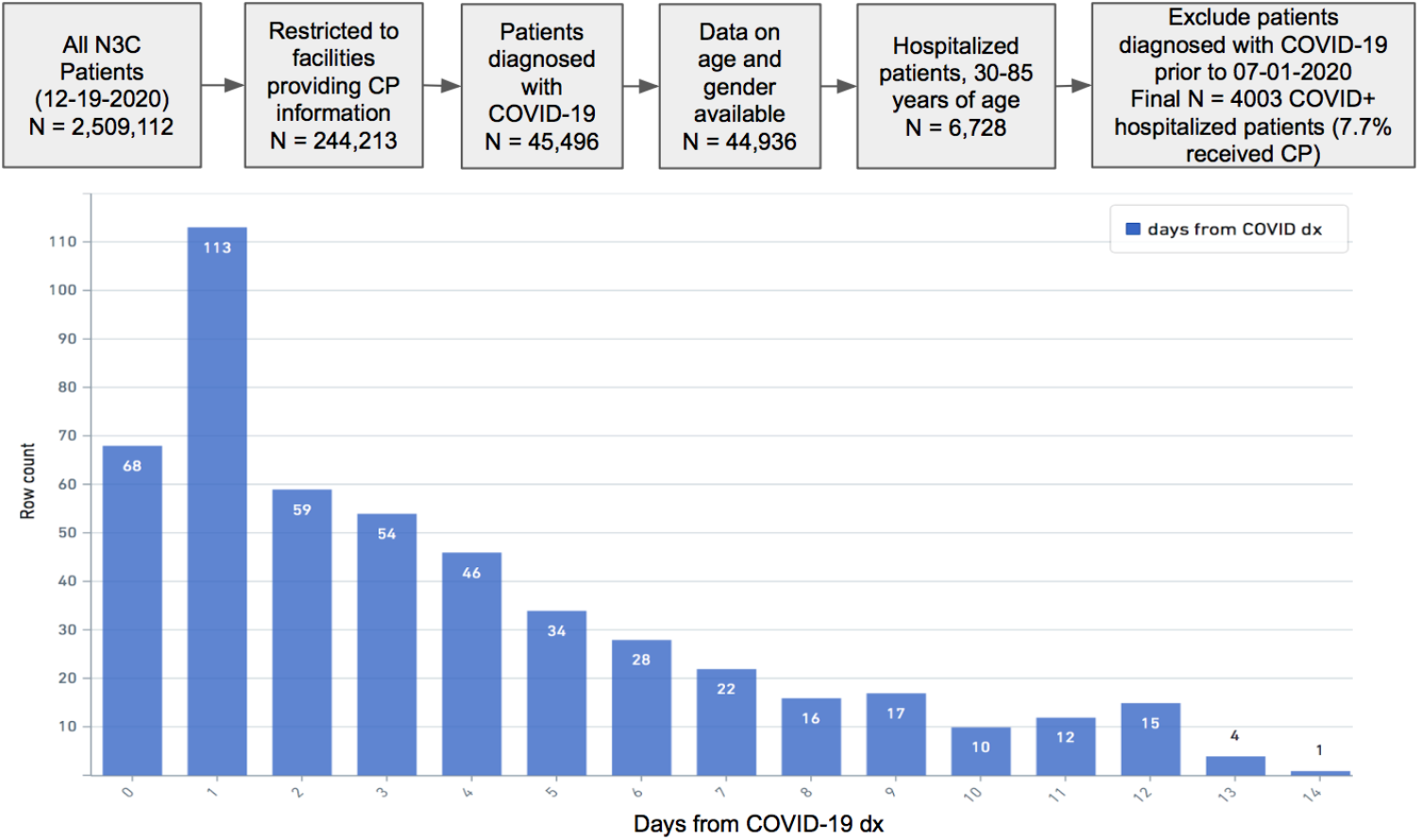
Data selection filter (top) and CP administration distribution from day of COVID-19+ diagnosis (bottom) in N3C (12-19-2020 release).

### Identification of Outcomes, Covariates and Treatment

We assessed live discharge from hospitalization, and thus hospitalization duration from treatment or in-hospital death as the two outcomes. Live discharge was determined through N3C *harmonized_visit* table which collates information from *visit_occurrence* table.^18^ In-hospital death was determined through the *death* table provided such death occurred during hospitalization.^24^ Covariates (i.e. predictors) were identified using OHDSI ^24,25^ concept sets. Treatment (CP administration) was identified by case insensitive string matching *TRANSFUSION*CONVALESCENT* *device_exposure* or *procedure_occurrence* tables.^19^ String matching was used as opposed to code sets because CP transfusion information was sent as free text as opposed to coded set by the subset of N3C data providers sending CP information.

### Statistical Analysis

We performed multi-predictor propensity score based cohort matching to account confounding in the compared cohorts.^26,27^ The 26 pre-COVID-19 diagnosis predictors used to construct a propensity score model for cohort matching were: gender, age, race, geographical region, Charlson Comorbidity Index (CCI)^28^ categories (0, 1, 2-3, 4-5, 6+), dexamethasone, specific CCI conditions (cancer, HIV, myocardial infarction, stroke, rheumatic disease, peripheral vascular disease, renal disease, congestive heart failure), prior medical disposition (smoker, diabetic, chronic respiratory disorder, hypertensive, hyper cholesterol), prior access to medical care (through prior monthly outpatient, inpatient and ER visits, medication, procedure, hospitalization rates), BMI and data provider. We additionally used 9 Day-of-CP-Administration predictors which included minimum systolic blood pressure, minimum Glomerular Filtration Rate (GFR), minimum O2 saturation, minimum bicarbonate, maximum respiratory rate, maximum heart rate, maximum temperature, maximum C-reactive protein (CRP) and maximum erythrocyte sedimentation rate (ESR) rate. We used a Bayesian logistic model^29^ to have numerically stable estimates. Patients who received CP were divided into subsets that received plasma only on Day 0, 1, 2, 3 and 4 and not prior to that specified day. For each subset a cohort was created using propensity scores via nearest neighbor matching with replacement (Figure 2).^27^ For each cohort we performed survival analysis ^30,31^ using Cumulative Incidence Function (CIF)^32^ and followed up with Gray’s CIF equality test.^33^ CIF estimates the incidence of the occurrence of an event (live discharge) while taking into account competing risk (death) while Gray’s test appropriately^34^ determines the significance of treatment on the competing events we analyzed.^30-16^ For each cohort we also performed cause specific hazard ratios^30^ at 95% confidence interval followed up with Cox’s test.^31^ A hazard ratio > 1.0 indicates a beneficial effect on risk of live hospital discharge. A hazard ratio of < 1.0 indicates a beneficial effect on risk of mortality. The outcome of interest for the survival analysis was live hospital discharge with death as a competing risk. We developed Java/Python/R routines to prepare and analyze N3C data. COVID-19 diagnosis was determined through non-immunoglobulin based COVID-19 tests.

**Figure 2:**
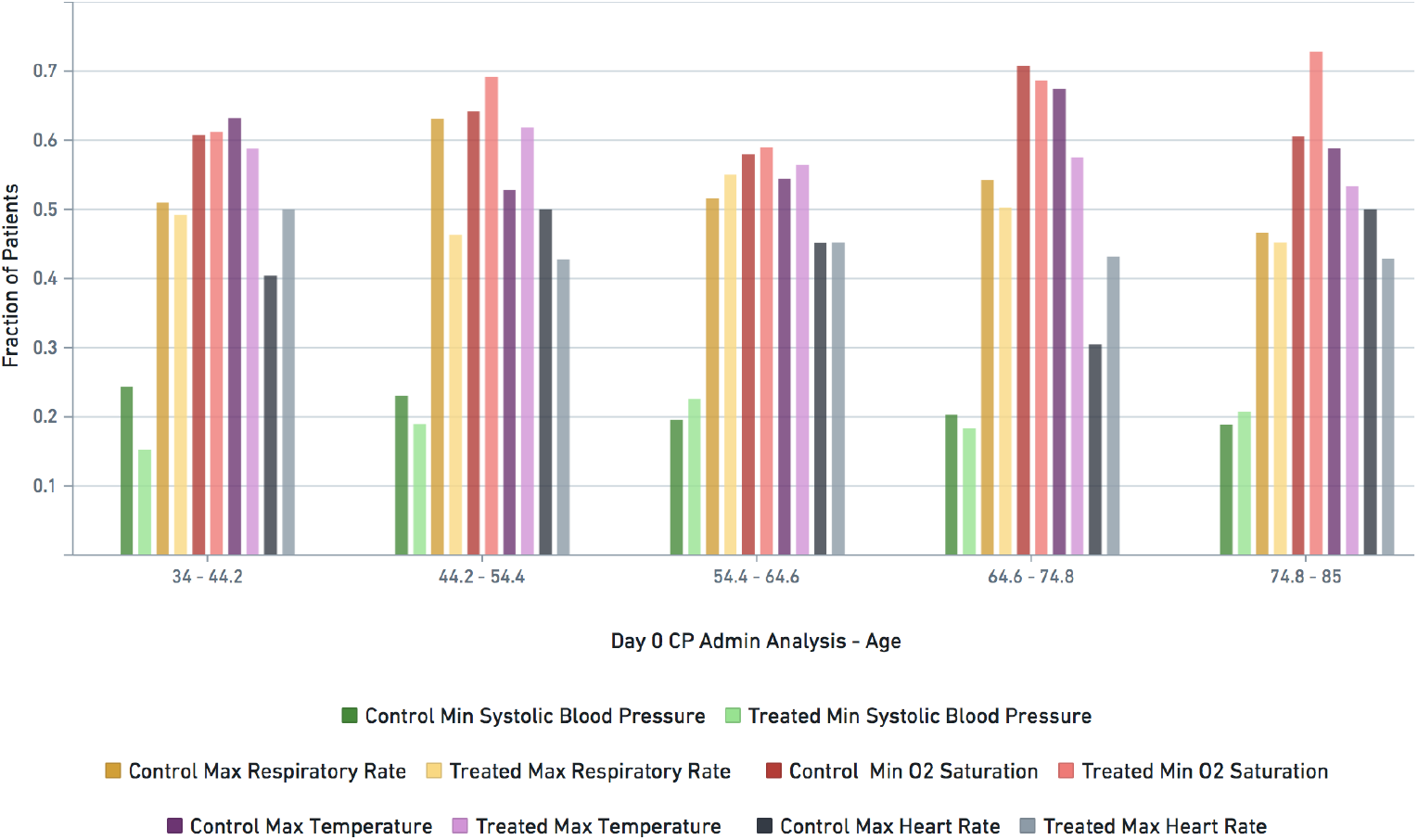
Patient state indicator distribution across age groups (as fraction of age group) for treated (with CP on Day 0) and matched control cohorts used for statistically significant clinical effect analysis. The darker shade of each colored pair is Control (untreated), while the lighter shade is Treated (with CP). For example, the dark purple (control cohort) and light purple (treated cohort) bars on each age group represent max temperature (normalized between 0 and 1).

Specific Day-of-CP-Administration predictors were used as they are general markers of patient state and have been shown to be important in prediction models for COVID-19 severity and in comparative effectiveness analyses for COVID-19 therapeutics. Minimum GFR was used as a marker for renal function, minimum O2 saturation and maximum respiratory rate were used as markers of respiratory disease. Bicarbonate was used as a marker for tissue perfusion.

Maximum ESR, maximum CRP, maximum temperature, and maximum heart rate were used as markers for inflammation.^35-38^ Follow-up was censored at 84 days.

## RESULTS

After excluding patients from providers that did not encode CP, that were age or gender, or were diagnosed with COVID-19 prior to July 1, 2020, there were 4,003 patients hospitalized with COVID-19, of whom 312 (7.7%) had received CP within 14 days of COVID-19 diagnosis (Figure 1).

In the above cohort, 197 (4.9%) received CP within Day 0, 1, 2, 3 or 4 of COVID-19 diagnosis. Multi-predictor propensity score matching based on both pre-COVID-19 predictors and

Day-of-CP-Administration predictors produced a matched comparison cohort for each treatment day. As a visual example, the predictor match for CP administered on day 0 is shown in Figure 2.

Table 1 shows the characteristics of the patients who received CP treatment and the matched cohorts for each treatment day. Overall, treated patients were predominately white (63%) and male (57%) with a mean age of 64. Of particular note is that these patients tended to be overweight or obese (mean BMI: 32kg/m^2); 44% were without prior comorbidities based on CCI, 46% were hypertensive, 37% diabetic and 12% with chronic respiratory issues. The matched cohort tracks these numbers closely overall *except* for day 4 which shows specifically smoker, diabetic, Charlson comorbidity predictor asymmetricity.

**Table 1:** **Characteristics of patients treated with convalescent plasma (CP) on (Day 0, 1, 2, 3 and 4) after COVID-19 diagnosis and propensity matched controls.**

| Day of CP Administration | Day 0 |  | Day 1 |  | Day 2 |  | Day 3 |  | Day 4 |  | Overall Cohort |  |
| --- | --- | --- | --- | --- | --- | --- | --- | --- | --- | --- | --- | --- |
| Characteristic | CP Treated | Matched Control | CP Treated | Matched Control | CP Treated | Matched Control | CP Treated | Matched Control | CP Treated | Matched Control | CP Treated | Matched Control |
| **Total N** | 48 | 48 | 69 | 69 | 25 | 25 | 27 | 27 | 28 | 28 | 197 | 197 |
| Deaths N (%) | 9<br>(18.75) | 15<br>(31.25) | 12<br>(17.39) | 10<br>(14.49) | 4<br>(16.0) | 2<br>(8.0) | 4<br>(14.81) | 6<br>(22.22) | 8<br>(28.57) | 2<br>(7.14) | 37<br>(18.78) | 35<br>(17.77) |
| Average Age (std) | 63.15<br>(12.64) | 62.55<br>(13.59) | 64.40<br>(13.54) | 63.13<br>(14.97) | 64.04<br>(14.23) | 65.84<br>(10.87) | 67.08<br>(9.59) | 68.45<br>(13.96) | 62.75<br>(15.31) | 60.65<br>(12.18) | 64.18<br>(13.15) | 63.77<br>(13.72) |
| Average BMI scale (kg/m<sup>2</sup>) | 0.549<br>(34.86) | 0.500<br>(33) | 0.493<br>(32.73) | 0.521<br>(33.79) | 0.377<br>(28.32) | 0.391<br>(28.85) | 0.507<br>(33.26) | 0.430<br>(30.34) | 0.455<br>(31.29) | 0.480<br>(32.24) | 0.482<br>(32.31) | 0.480<br>(32.24) |
| Race |  |  |  |  |  |  |  |  |  |  |  |  |
| White (%) | 62.5 | 72.92 | 68.12 | 63.77 | 60 | 76 | 62.96 | 70.37 | 57.14 | 35.71 | 63.45 | 64.47 |
| African American (%) | 20.83 | 10.42 | 13.04 | 18.84 | 8 | 8 | 11.11 | 11.11 | 17.86 | 21.43 | 14.72 | 14.72 |
| Other (%) | 10.42 | 10.42 | 13.04 | 10.14 | 20 | 12 | 22.22 | 18.52 | 21.43 | 35.71 | 15.74 | 15.23 |
| Missing (%) | 6.25 | 6.25 | 5.8 | 7.25 | 12 | 4 | 3.7 | 0 | 3.57 | 7.14 | 6.09 | 5.58 |
| Average CCI (std) | 1.94<br>(2.65) | 2.10<br>(3.28) | 1.86<br>(2.55) | 1.26<br>(2.37) | 2.72<br>(2.78) | 3.00<br>(2.36) | 1.59<br>(2.36) | 1.85<br>(2.28) | 2.71<br>(3.72) | 2.04<br>(3.53) | 2.07<br>(2.77) | 1.88<br>(2.82) |
| CCI Category |  |  |  |  |  |  |  |  |  |  |  |  |
| A (0) (%) | 47.92 | 54.17 | 47.83 | 62.32 | 28 | 20 | 44.44 | 40.74 | 42.86 | 53.57 | 44.16 | 50.76 |
| B (1) (%) | 14.58 | 4.17 | 13.04 | 11.59 | 12 | 8 | 22.22 | 22.22 | 10.71 | 14.29 | 14.21 | 11.17 |
| C (2-3) (%) | 12.5 | 16.67 | 18.84 | 13.04 | 32 | 36 | 18.52 | 14.81 | 17.86 | 10.71 | 18.78 | 16.75 |
| D (4-5) (%) | 10.42 | 14.58 | 5.8 | 7.25 | 12 | 12 | 7.41 | 14.81 | 7.14 | 7.14 | 8.12 | 10.66 |
| E (6+) (%) | 14.58 | 10.42 | 14.49 | 5.8 | 16 | 24 | 7.41 | 7.41 | 21.43 | 14.29 | 14.72 | 10.66 |
| Diabetic (%) | 25 | 18.75 | 34.78 | 23.19 | 44 | 48 | 48.15 | 40.74 | 46.43 | 32.14 | 37.06 | 28.93 |
| Smoker (%) | 6.25 | 6.25 | 8.7 | 8.7 | 4 | 4 | 0 | 0 | 14.29 | 0 | 7.11 | 5.08 |
| Female (%) | 41.6 | 39.58 | 44.93 | 46.38 | 44 | 52 | 40.74 | 40.74 | 42.86 | 35.71 | 43.15 | 43.15 |
| Hypertensive (%) | 37.5 | 29.17 | 39.13 | 28.99 | 56 | 68 | 55.56 | 62.96 | 64.29 | 53.57 | 46.7 | 42.13 |
| Elevated<br>Cholesterol (%) | 8.33 | 6.25 | 10.14 | 2.9 | 16 | 28 | 7.41 | 11.11 | 7.14 | 14.29 | 9.64 | 9.64 |
| Chronic<br>Respiratory Issue<br>(%) | 18.75 | 16.67 | 13.04 | 7.25 | 12 | 20 | 3.7 | 0 | 10.71 | 7.14 | 12.69 | 10.15 |

Note: BMI described as kg/m<sup>2</sup> and also rescaled to be between 0 and 1. Overall Cohort summary (two columns from the right) provided for reference only as study analysis aim was focused on specific days of CP administration.

For each of Day 0, 1, 2, 3 and 4, we *failed* to detect any statistically significant reduction in hospitalization duration through both Gray’s CIF equality test and Cox’s cause-specific hazard ratio tests (Table 2 and Figure 3). For the CP-administered on day 0, the cause-specific death hazard ratio was 0.45 but this association was not significant (p-value 0.06). For the CP-administered on day 4, there was a statistically significant *increased* death hazard ratio (Table 2 and Figure 3) but as the cohort size (N=28) was low for the day 4 analysis, and as smoker, diabetic, Charlson comorbidity matching were asymmetric, we are unable to come to a definitive conclusion for day 4.

**Table 2:** Gray’s equality test on CIF, Cause Specific Hazard Ratio and associated Cox regression p-Value for CP administration on specific days after non-Ig test based COVID-19 diagnosis. For example, the table shows that for CP administered on Day 0 the CIF did not differ for hospitalization duration or death analysis (Gray’s Test is not <= 0.05) and additionally while the *odds of discharge* and *odds of death* were both lower (Cause Specific Hazard Ratio column) neither were statistically significant (Cox regression is not <= 0.05).

| Analysis | Event | Gray's Test p-Value | Cause Specific Hazard Ratio | 95% Confidence Intervals | Cox Regression p-Value |
| --- | --- | --- | --- | --- | --- |
| CP @ Day 0 | Live Discharge | 0.77 | 0.69 | 0.43-1.11 | 0.13 |
| CP @ Day 0 | Death | 0.12 | 0.45 | 0.19-1.04 | 0.06 |
| CP @ Day 1 | Live Discharge | 0.34 | 0.74 | 0.51-1.07 | 0.11 |
| CP @ Day 1 | Death | 0.75 | 0.88 | 0.38-2.04 | 0.76 |
| CP @ Day 2 | Live Discharge | 0.48 | 0.94 | 0.51-1.75 | 0.85 |
| CP @ Day 2 | Death | 0.37 | 1.69 | 0.31-9.25 | 0.55 |
| CP @ Day 3 | Live Discharge | 0.73 | 0.86 | 0.46-1.60 | 0.63 |
| CP @ Day 3 | Death | 0.52 | 0.58 | 0.16-2.10 | 0.41 |
| CP @ Day 4 | Live Discharge | 0.64 | 1.37 | 0.70-2.70 | 0.36 |
| CP @ Day 4 | Death | 0.047 | 5.23 | 1.08-25.31 | 0.04 |

**Figure 3:**
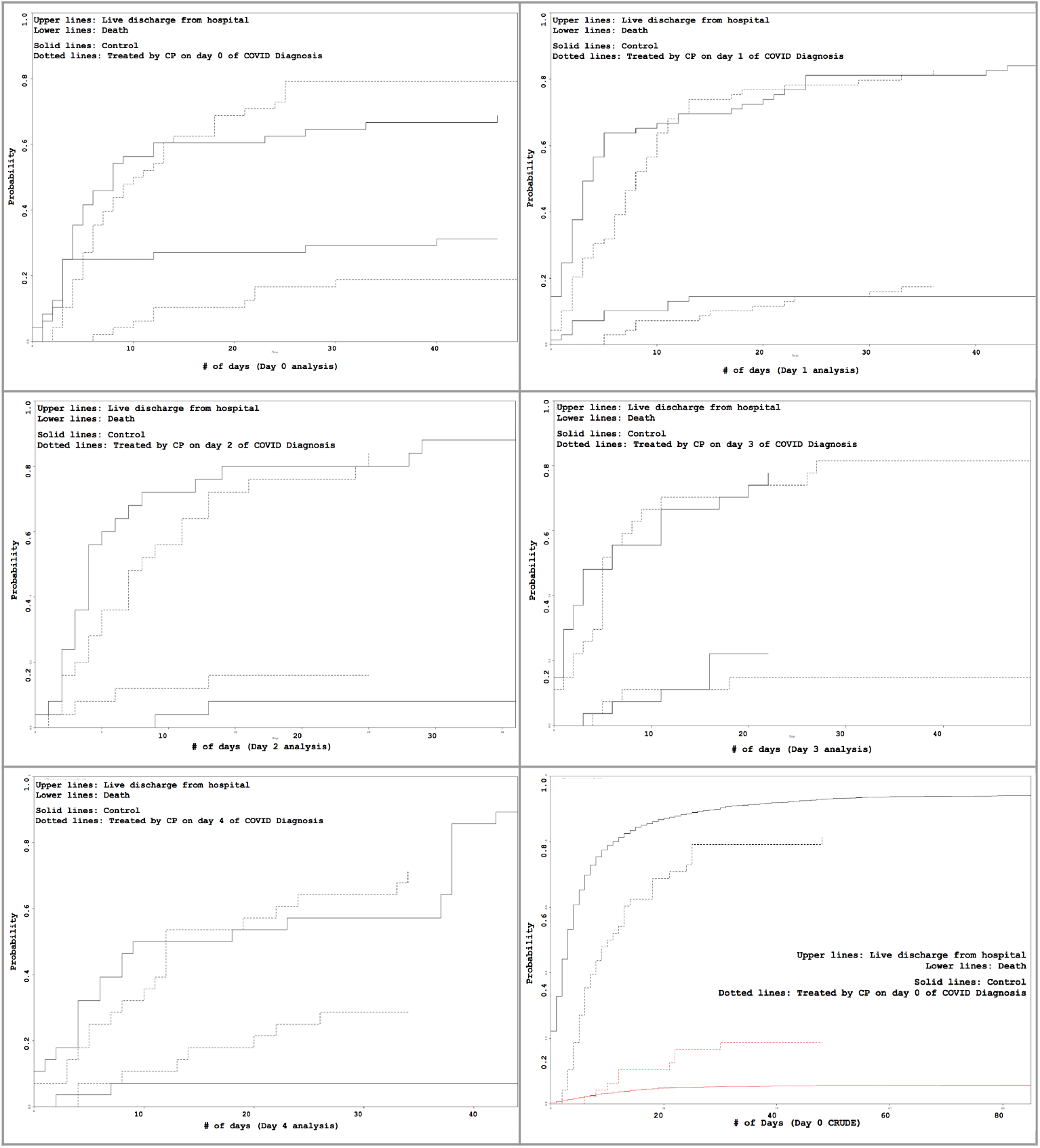
CP Administration on Day X (X=0,1,2,3,4) CIF survival visualization with live hospital discharge duration as the primary endpoint and death as competing risk. Bottom right: CRUDE analysis *without* matching for day 0 CP treatment.

## DISCUSSION

In this study, treatment with CP on Day 0, 1, 2, 3 and 4 of COVID-19 diagnosis demonstrated no significant effect on duration of hospitalization among COVID-19 patients. In addition to duration of hospitalization as determined by live discharge from the hospital, in-hospital deaths were also assessed in this study which also showed no association with CP treatment.

Assessment of the effectiveness of CP administration has shown mixed results across RCT and observational studies as summarized in Table 3. When comparing the results of our study with those of prior studies, following key points were observed:

**Table 3:** **Existing Randomized Controlled Trials (RCT) and Observational Studies on CP Administration Effectiveness as COVID-19 Treatment. The Last Row Summarizes the Results from this (Alamgir et al) N3C Study.**

| Author | Type of Study | N (% treated) | Administration Time Frame (from hospitalization unless otherwise noted) | Pandemic Timeframe | Titer Stratification | Country | Conclusion |
| --- | --- | --- | --- | --- | --- | --- | --- |
| Joyner et al (4) | Observational (no matching) | 3,082 (26.9%) | <= 72 hours | Apr 4 2020 - Jul 4 2020 | Yes | USA | Among patients hospitalized with Covid-19 who were not receiving mechanical ventilation, transfusion of plasma with higher anti-SARS-CoV-2 IgG antibody levels was associated with a *lower risk of death* than transfusion of plasma with lower antibody levels. |
| Simonovich et al (5) | RCT | 333 (68.4%) | Avg 8 (5 - 10) days from symptom onset | May 28 2020 - Aug 27 2020 | N/A | Argentina | *No significant differences* were observed in clinical status or overall mortality between patients treated with convalescent plasma and those who received placebo. |
| Libster et al (6) | RCT | 160 (50%) | <= 72 hours | Jun 4 2020 - Oct 25 2020 | N/A | Argentina | Early administration of high-titer convalescent plasma against SARS-CoV-2 to mildly ill infected older adults *reduced the progression of Covid-19* (severe respiratory disease endpoint). |
| Horby et al (7) | RCT | 11,199 (48.54%) | Avg 9 (6 - 12) days from symptom onset | May 28 2020 - Jan 15 2021 | N/A | United Kingdom | Among patients hospitalised with COVID-19, high-titer convalescent plasma *did not improve survival or other prespecified clinical outcomes*. |
| Agarwal et al (8) | RCT | 464 (50.64%) | N/A | Apr 22 2020 - July 14 2020 | N/A | India | Convalescent plasma was *not associated with a reduction in progression to severe covid-19* or all cause mortality |
| Li et al (9) | RCT | 103 (50.4%) | <= 72 hours | Feb 14 2020 - Apr 1 2020 | N/A | China | Among patients with severe or life-threatening COVID-19, convalescent plasma therapy added to standard treatment *did not significantly improve the time to clinical improvement* within 28 days, although the trial was terminated early and may have been underpowered to detect a clinically important difference. |
| Janiaud et al (10) | Review and Meta-analysis | 1,060 (56.13%) from peer reviewed RCTs only. | N/A | Feb 14 2020 - Oct 25 2020 (peer reviewed RCTs only) | N/A | India, Argentina, Bahrain, China, The Netherlands, Spain, UK | Treatment with convalescent plasma compared with placebo or standard of care was *not significantly associated* with a decrease in all-cause mortality or with any benefit for other clinical outcomes. |
| Rogers et al (11) | Observational (with matching) | 241 (36.15%) | Avg 7 days from symptom onset | Pre May 31 2020 | N/A | USA | Treatment of severe COVID-19 with convalescent plasma *was not associated with a significantly different risk of in-hospital mortality or rate of hospital discharge* as compared to standard of care |
| Sostin et al (12) | Observational (with matching) | 96 (32.29%) | 10 (7–13) days from onsite, and 3 (2 - 5) days from hospitalization | Apr 10 2020 - May 5 2020 | N/A | USA | Convalescent plasma *did not reduce in-hospital mortality in our sample, nor did it reduce length of stay.* |
| Salazar et al (13) | Observational (with matching) | 387 (35.14%) | <= 72 hours and > 72 hours (two groups) | March 28 2020 - July 6 2020 |  | USA | The data suggest that transfusion of high anti-RBD IgG titer convalescent plasma early in the hospital course significantly *reduces mortality* |
| Liu et al (14) | Observational (with matching) | 195 (20%) | 4 days (0-7 days) | Mar 24 2020 - Apr 8 2020 | Yes | USA | Survival *improved* in plasma recipients |
| Gharbharan (15) | RCT | 86 (50%) | 2 days (1-3 days) | Apr 8 2020 - N/A | No | The Netherlands | Most COVID-19 patients already have high neutralizing antibody titers at hospital admission. *No difference* in mortality, hospital stay was observed between plasma treated patients and patients on standard of care. Study terminated early. |
| Budhiraja et al (16) | Observational (no matching) | 694 (47.98%) | N/A | May 1 2020 - Aug 31 2020 | Yes | India | In patients with COVID-19 admitted to ICU, *mortality was significantly lower in plasma arm*. This benefit of reduced mortality was most seen in age group 60 to 74 years driven mostly by females of this age group. Significant difference in mortality was observed in patients with one comorbidity. Moreover, *patients on ventilator had significantly lower mortality* in the plasma arm; particularly so for patients on invasive mechanical ventilation |
| Alamgir et al (This Study) | Observational (with matching) | 394 (50%) | Specifically (Day 0,1,2,3,4) from COVID-19 diagnosis analysis | July 1, 2020 - December 19, 2020 | No | USA | *No significant differences* found in hospitalization duration from treatment or in-hospital mortality as a result of CP administration on day 0,1,2,3 or 4th day from COVID-19 diagnosis. Day 0 CP administration showed a *non-statistically significant effect* on reduced mortality odds. |

- Findings are *consistent* with 5 of the RCTs of Table 3 (Simonovich et al, Horby et al, Agarwal et al, Li et al, Gharbaran et al)
- Findings are *inconsistent* with 1 of the RCTs (Libster et al).
- Findings are *consistent* with two of the observational studies that performed patient matching (Rogers et al, Sostin et al)
- Findings are *inconsistent* with two of the observational studies that performed patient (Salazar et al, Liu et al).
- Findings are *inconsistent* with the only observational studies that did *not* perform patient matching (Budhiraja et al)
- The Joyner et al study is not comparable as it did not compare effect of CP administration with a control group that did not receive CP

The studies with which our results are inconsistent can be further de-constructed. The Budhiraja et al study reported significant positive outcomes for patients admitted to ICU and administered CP. This ICU+CP treated cohort had 171 people over 60 years of age compared to 184 people over 60 years of age in the control cohort and a significant difference given effect of age on COVID-19 mortality.^39^ Additionally, the same study had 30% less people with 3+ co-morbidities in the ICU+CP treated group compared to control group which is also a significant difference given the effect of multiple co-morbidities in COVID-19 mortality.^40^ In Salazar et al and Liu et al the number of patients with CP included in the statistical analysis was 143 and 39, which is 27% and 80% *lower* than the number of patients with CP that was analyzed in our study. Finally, the Libster et al RCT was reasonably comparable between treated and untreated groups except for age where the number of people over 75 years old was 20% higher in the untreated group (N=48) as compared to the treated group (N=40) - a significant difference given effect of age on COVID-19 disease progression and mortality.^39^

Stratified analysis of effect of CP administration on specific days on or after COVID-19 diagnosis and the use of markers for inflammation, respiratory distress and renal function on specific CP administration days employed in this study provide an objective view into efficacy of CP treatment on duration of COVID-19 related hospitalization.

### Limitations

Administration of CP was not consistently coded in the N3C data. To restrict our analysis to patients who had received CP rather than other plasma products, we excluded all data from providers without specific codes or descriptions for CP data. That restriction reduced our potential population by over 90%. While we attempted to match patient severity using 9 day of CP predictors based on data available within N3C, there could be other predictors that better represent patient status for matching purposes. Furthermore, the study could be augmented, pending data availability, with information on the oxygen device that was used during the treatment period. Another limitation is that the N3C dataset did not track whether a patient was involved in a COVID-19 vaccination trial which may skew results. This, however, is unlikely as vaccinated individuals are unlikely to be hospitalized for COVID-19 given the timeframe this analysis considered is between 07/01/2020 and 12/19/2020. Finally, while the research team considered use of titer and blood type, the analyzed N3C dataset did not contain titer levels of CP that was administered and was sparse in blood type information.

### Future work

Our data do not demonstrate any clear benefit for CP administration in hospitalized patients with COVID-19 within the first 4 days of hospitalization. There was a trend towards reduced mortality for CP administered on Day 0 but a potential for increased mortality if administered on day 4. More work needs to be done to understand the possibility that earlier treatment may have a beneficial effect on COVID-19 outcomes, particularly as decades ago research pointed to similar observations^41^, versus the possibility that later treatment may be harmful.

## Data Availability

National Institute of Health's (NIH) National COVID Cohort Collaborative (N3C) has clear procedures for researchers to gain access to the data (1000+ researchers already have access to the data) and as such the patient statistical analysis of this manuscript is transparent and repeatable. https://ncats.nih.gov/n3c provides data access request process details. Supplementary materials to this manuscript provided at https://www.ariscience.org/p3_sc2_paper_03.html include R analysis code.

## ACKNOWLEDGEMENTS

The statistical analyses we performed were conducted with data or tools accessed through the NCATS N3C Data Enclave and supported by NCATS U24 TR002306. This research was possible because of the patients whose information is included within the data and the organizations and scientists who have contributed to the on-going development of this community resource.^17^ This work was supported by the Intramural Research Program of the National Institutes of Health, National Cancer Institute, Division of Cancer Epidemiology and Genetics.The content of this publication and the opinions expressed do not necessarily reflect the views or policies of the Department of Health and Human Services, the National Institutes of Health or the National Cancer Institute nor does mention of trade names, commercial products, or organizations imply endorsement by the U.S. Government. We also acknowledge support from the N3C Publication Committee in the preparation of this manuscript.

## AUTHOR CONTRIBUTIONS

JA, SS, PK, XC wrote the first draft of the manuscript. JA, SS, MY, BG, RE, HM, CA, XC set up the statistical test design. RA, NM, BG, SB, PK, TO provided clinical input and patient severity predictor considerations and use thereof. XC wrote R code that was reviewed by JA, MY, SS. JA, SH, XC wrote data preparation code. TO, PK, CA, SB, VM, JA provided existing CP literature support. SB spurred the initial idea to look at CP within N3C. All authors reviewed and edited the manuscript. JA, XC, RE prepared tables and figures with input from other authors.

Authorship was determined using ICMJE recommendations.

## PATIENT CONSENT FOR PUBLICATION

Not Applicable

## ETHICS APPROVAL AND CONSENT TO PARTICIPATE

National Institute of Health’s (NIH) National COVID Cohort Collaborative (N3C) Data Utilization Request Approval committee approved the data utilization request of this project (RP-C80011)

**Supplemental Appendix** (including all R code used) is available at: https://www.ariscience.org/p3_sc2_paper_03.html

